# Lack of sufficient public space can limit the effectiveness of COVID-19’s social distancing measures

**DOI:** 10.1101/2020.06.07.20124982

**Authors:** Alexandre Nobajas, Joan Ganau i Casas, Daniel Paül i Agustí, Adam Peacock

## Abstract

One of the primary strategies of slowing down the COVID-19 pandemic has been the establishment of social distancing rules that recommend keeping a buffer distance between individuals, and this has proven effective in helping in reducing the basic reproduction number [R _0_]^1^. However, social distancing rules have put the use of public spaces in densely populated places under strain, and this is especially important as some of the most virulent outbreaks of the COVID-19 pandemic have been in compact cities. It is therefore fundamental to take into account each neighbourhood’s morphological characteristics and the potential population densities each street, square or park can accommodate under such new regulations in order to effectively enforce social distancing rules. Otherwise, certain areas may be rapidly overwhelmed by crowds with citizens unable to maintain the minimum safe distance between individuals. In this paper, we develop a method to identify the potential public space accessibility if social distancing rules are followed and we apply it to three global and highly affected by COVID-19 cities. Our research finds that, at micro level there are important inequalities between neighbourhoods, so people will struggle to comply with social distancing rules and consequently it will make controlling infection rates more difficult.

---

The COVID-19 pandemic has altered perceptions and uses of public spaces globally^2^. Citizens have grappled with not being able to use public spaces to meet with neighbours, friends or family^3^. For some, gatherings in public areas have become viewed as dangerous and are avoided due to the risk of coming into contact with those who might be infected^4^. Yet, as cities continue to adapt to the outbreak, public spaces are likely to become more utilized once again. In this context, public space availability is key, but the urban fabric of cities can dramatically vary, with the quantity and quality of public spaces often related to each neighbourhood’s wealth^5^, a factor which can deepen urban segregation and fragmentation^6^. Subsequently, a one rule fits all solution is unlikely to work effectively across all neighbourhoods. Each city will need to manage its respective public spaces independently and in detail in order to ensure that social distancing rules are adhered to, whilst simultaneously allowing minimal use of public space for daily activities^7^.

The field of urban data applications provides the tools necessary to help accommodate such considerations, by allowing planners and policy makers to quantify and model how many people can access each public space while keeping a safe distance during a pandemic outbreak. This is something that has not been researched enough^8^, primarily due to the unprecedented scale of the COVID-19 crisis. Therefore, this communication focuses in accurately measuring the maximum capacity of public space at a microscale level. Without this information, it may be impossible for citizens to effectively follow the regulations and for regulators to enforce them. Urban data applications, coupled with geographical information science, prove excellent tools in mapping potential public space population densities and identifying which areas are more likely to exceed their maximum capacity if social distancing rules are followed.

## Public space density and COVID-19 policy

By using publicly available data at the highest spatial resolution and granularity available, the Spanish rules for social distancing for Madrid and Barcelona, two of the worst affected cities in Europe with 37,097 and 17,752 detected cases respectively (30 May 2020)^9^, can be modelled and mapped. Comparatively, the same can be accomplished in New York City, which has 201,806 confirmed cases (4 June 2020)^10^. Local regulations in Spain establish that where there is a risk of COVID-19 infection spreading, citizens cannot go further than 1km around their place of residence and that they must keep at least 2 meters apart^11^, so each individual needs a minimum of 3.14m^2^ -πr^2^-of personal space in order to comply. If blanket regulations at nationwide level are used, for example in Barcelona, the city has enough public space to hold almost 17M people within its limits. When rules are mapped, however, it becomes clear that the current broad regulations are not detailed enough to successfully manage public space use in a manner that guarantees a safe and equal access. Figure 1 indicates how many people have access to each social distancing unit of 3.14m^2^ of public space and could therefore potentially visit that spot at any given time. Figure 2 uses the same method adapted to New York’s social distancing rule of 6ft^12^.

**Figure 1.**
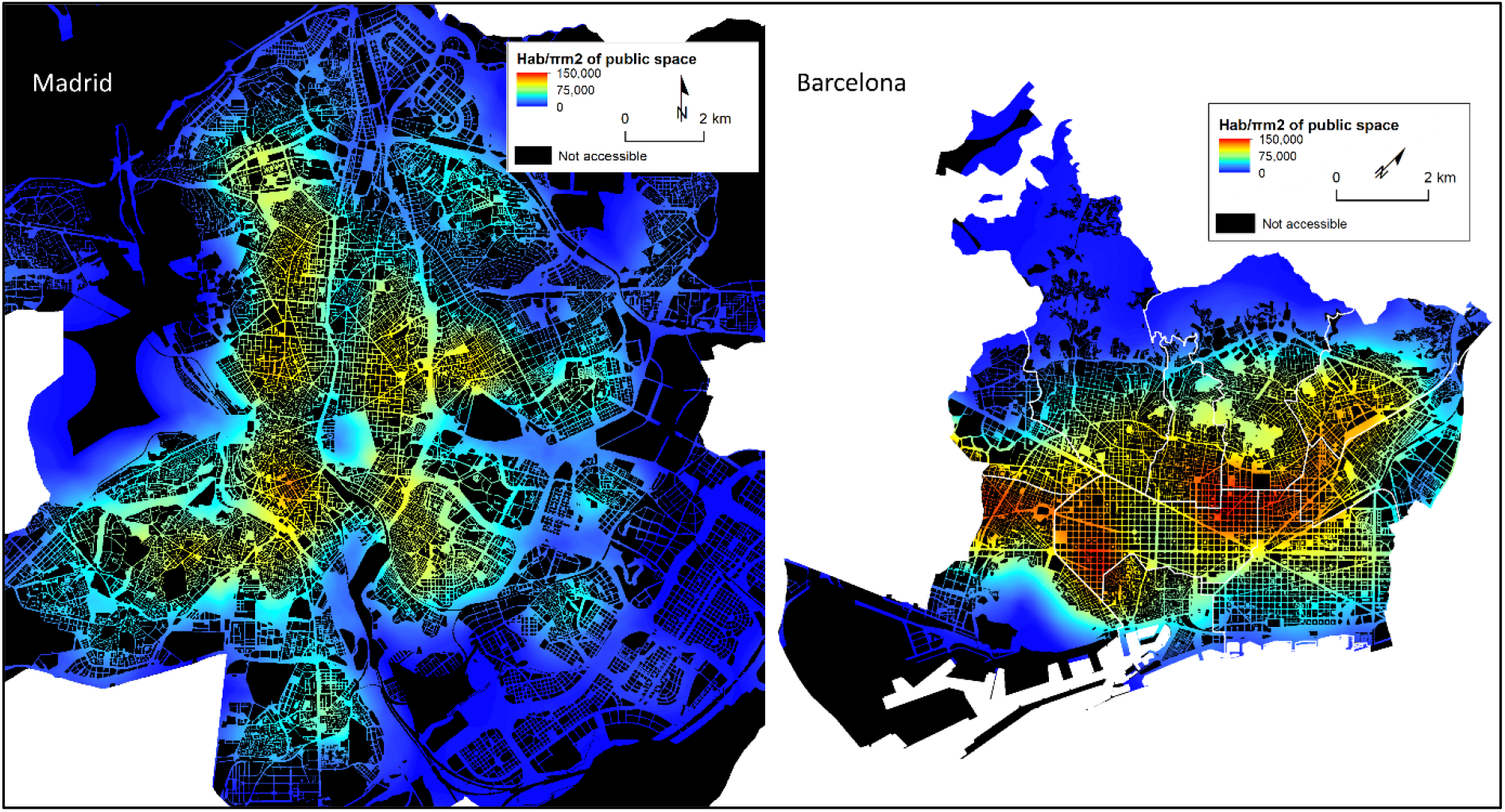
Potential public space population density in Madrid and Barcelona if people are to keep 2m apart and space is accessible to people residing 1km from it.

**Figure 2.**
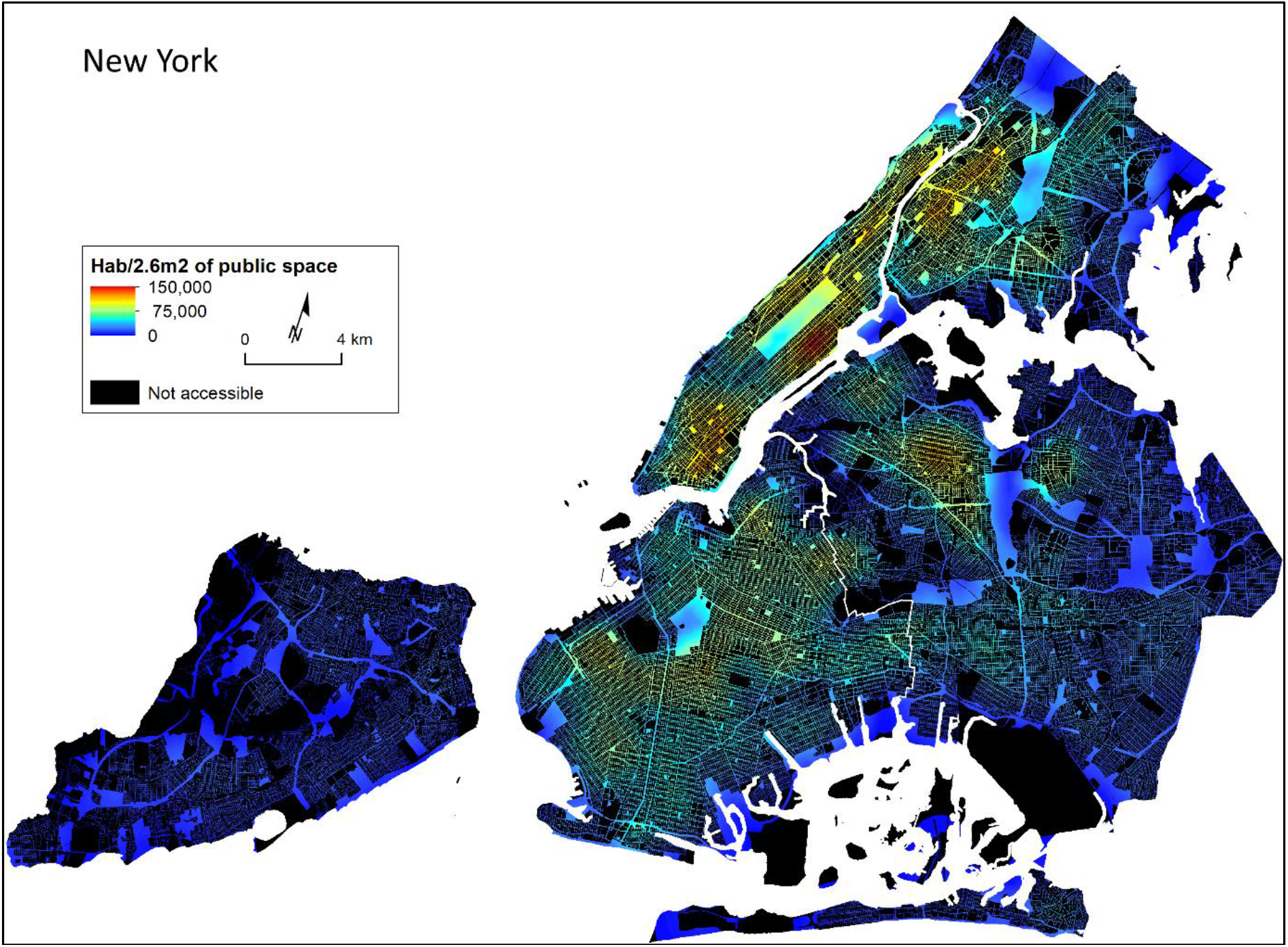
Potential public space population density in New York if people are to keep 6ft apart. Population has been calculated by how many people reside within 1km of each social distancing unit of public space.

Results are a theoretical maximum public space population density as the model assumes that all public space is available for pedestrian use. From this baseline, planners need to consider that any other uses, such as on-street parking, restaurant terraces, road traffic or bans in the use of parks, will further decrease the availability of public space, thus exacerbating its scarcity. Moreover, since such spaces are a limited resource, public space and urban nature need to be managed very carefully in order to limit the spread of the disease and, similarly, in order to minimise social inequalities^13^. While under social distancing rules certain areas of a city may not have to modify their mobility patterns, other zones will need to be micromanaged in order for all citizens to be able to safely exit their home. Otherwise only those who are fortunate enough to live in an area where public space density is low will be able to safely go outside. Initiatives such as reducing the space dedicated to public transport or setting a schedule so not everybody is allowed to go out at once are, although often unwelcome, necessary initiatives that do not need to be applied everywhere, just in those areas where geospatial analysis highlights the need to.

## Conclusion

Interpersonal pace is a key factor in the spread and control of COVID-19, and consequently most countries have established regulations to enforce social distancing between individuals. This paper shows that most major urban areas are not adequately prepared to respond to such an infectious disease, as in certain areas public space can become quickly overwhelmed simply by locals exiting their homes due to an inadequate provision of available public space. Furthermore, competing uses of such spaces mean that not all public space is available for pedestrians to use as they desire, exacerbating the issue. The current pandemic therefore poses two challenges, an immediate one, as public space will need to be micromanaged in order to prevent the spread of the disease and longer term one, as future city planning will need to take potential future pandemics into account. In this sense the COVID-19 pandemic has proven that medical recommendations alone are not enough to prevent the spread of an illness and that space matters. This paper offers a useful methodological tool which helps address both of these challenges and puts social distancing requirements in the center of current and future morphological considerations.

## Methods

### Data and Analysis

Data to produce Figure 1 and 2 are from open and official sources. Land use for Barcelona (https://opendata-ajuntament.barcelona.cat/data/ca/dataset/mapa-usos-sol-wms) and Madrid (https://datos.madrid.es/portal/site/egob/menuitem.c05c1f754a33a9fbe4b2e4b284f1a5a0/?vgnextoid=81f9a6cb58f1c510VgnVCM2000001f4a900aRCRD&vgnextchannel=374512b9ace9f310VgnVCM100000171f5a0aRCRD&vgnextfmt=default) come from their respective city councils’ data services and refer to the year 2016. Land use data for the cities’ neighbouring areas comes from the Copernicus Land Monitoring Service (https://centrodedescargas.cnig.es/CentroDescargas/index.jsp#) and refer to the year 2012. Demographic data (https://www.ine.es/jaxiT3/files/t/es/xlsx/30832.xlsx?nocab=1) are from Spain’s National Statistical Institute and refers to the year 2017, while Census areas geographies refer to the year 2011 (https://www.ine.es/censos2011_datos/cartografia_censo2011_nacional.zip). For New York, data population data came from the census (https://www.census.gov/geographies/mapping-files/2010/geo/tiger-data.html) and land use from the council (https://www1.nyc.gov/assets/planning/download/zip/data-maps/open-data/nyc_mappluto_20v3_unclipped_shp.zip).

Public space was defined as all those places classed as streets and roads, parks and green spaces and not privately owned. The map’s resolution is 3.1415m^2^ (2.63m^2^ for NYC) per pixel, the size of a 1m (3ft for NYC) radius in order to reflect social distancing rules. Population distribution is obtained by the smallest census unit available, and that population is then equally distributed across the areas of all residential dwellings of the area. Potential public space density is calculated by counting how many people live in a 1km radius around each public space pixel and therefore can reach it. Data analysis was done in R, geographical analysis was done in QGIS and data visualisation was done using ArcMap.

### Limitations

Land use data for all three cities have small inaccuracies that while not being critical to the argument can be spotted by locals and those who know the city well, such as semi-private parks or open spaces that are not classed as public space.

## Data Availability

All data is freely available online and all links are included as part of the manuscript

https://opendata-ajuntament.barcelona.cat/data/ca/dataset/mapa-usos-sol-wms

https://datos.madrid.es/portal/site/egob/menuitem.c05c1f754a33a9fbe4b2e4b284f1a5a0/?vgnextoid=81f9a6cb58f1c510VgnVCM2000001f4a900aRCRD&vgnextchannel=374512b9ace9f310VgnVCM100000171f5a0aRCRD&vgnextfmt=default

https://centrodedescargas.cnig.es/CentroDescargas/index.jsp#

https://www.ine.es/censos2011_datos/cartografia_censo2011_nacional.zip

https://www.census.gov/geographies/mapping-files/2010/geo/tiger-data.html

https://www1.nyc.gov/assets/planning/download/zip/data-maps/open-data/nyc_mappluto_20v3_unclipped_shp.zip

## Notes

### Competing Interest Statement

The authors have declared no competing interest.

### Funding Statement

No funding was received

### Author Declarations

Data utilized was from publicly available sources with no individual level identifiers. The authors self-exempted the data as not qualifying as human subjects research requiring IRB oversight.

